# A functional variant of the *SIDT2* gene involved in cholesterol transport is associated with HDL-C levels and premature coronary artery disease

**DOI:** 10.1101/2020.09.19.20197673

**Authors:** Paola León-Mimila, Hugo Villamil-Ramírez, Luis R. Macias-Kauffer, Leonor Jacobo-Albavera, Blanca E. López-Contreras, Rosalinda Posadas-Sánchez, Carlos Posadas-Romero, Sandra Romero-Hidalgo, Sofía Morán-Ramos, Mayra Domínguez-Pérez, Marisol Olivares-Arevalo, Priscilla Lopez-Montoya, Roberto Nieto-Guerra, Víctor Acuña-Alonzo, Gastón Macín-Pérez, Rodrigo Barquera-Lozano, Blanca E. del Río-Navarro, Israel González-González, Francisco Campos-Pérez, Francisco Gómez-Pérez, Victor J. Valdés, Alicia Sampieri, Juan G. Reyes-García, Miriam del C. Carrasco-Portugal, Francisco J. Flores-Murrieta, Carlos A. Aguilar-Salinas, Gilberto Vargas-Alarcón, Diana Shih, Peter J. Meikle, Anna C. Calkin, Brian G. Drew, Luis Vaca, Aldons J. Lusis, Adriana Huertas-Vazquez, Teresa Villarreal-Molina, Samuel Canizales-Quinteros

**Author notes:** Correspondence (T.V.-M.), (S.C.-Q.). These authors contributed equally to this work (Paola León-Mimila, Hugo Villamil-Ramírez, Luis R. Macias-Kauffer, Leonor Jacobo-Albavera).

## Abstract

Low HDL-C is the most frequent dyslipidemia in Mexicans, but few studies have examined the underlying genetic basis. Moreover, few lipid-associated variants have been tested for coronary artery disease (CAD) in Hispanic populations. Here, we performed a GWAS for HDL-C levels in 2,183 Mexican individuals, identifying 7 *loci*, including three with genome-wide significance and containing the candidate genes *CETP, ABCA1* and *SIDT2*. The *SIDT2* missense Val636Ile variant was associated with HDL-C levels for the first time, and this association was replicated in 3 independent cohorts (*P*=5.5×10^−21^ in the conjoint analysis). The *SIDT2*/Val636Ile variant is more frequent in Native American and derived populations than in other ethnic groups. This variant was also associated with increased ApoA1 and glycerophospholipid serum levels, decreased LDL-C and ApoB levels and a lower risk of premature CAD. Because SIDT2 was previously identified as a protein involved in sterol transport, we tested whether the SIDT2/Ile636 protein affected this function using an *in vitro* site-directed mutagenesis approach. The SIDT2/Ile636 protein showed increased uptake of the cholesterol analog dehydroergosterol, suggesting this variant is functional. Finally, liver transcriptome data from humans and the Hybrid Mouse Diversity Panel (HMDP) are consistent with the involvement of *SIDT2* in lipid and lipoprotein metabolism. In conclusion, this is the first study assessing genetic variants contributing to HDL-C levels and coronary artery disease in the Mexican population. Our findings provide new insight into the genetic architecture of HDL-C and highlight *SIDT2* as a new player in cholesterol and lipoprotein metabolism in humans.

## INTRODUCTION

Observational epidemiologic studies have reported that low plasma high density lipoprotein cholesterol (HDL-C) concentrations are an independent risk factor for cardiovascular disease.^1-4^ Heritability of HDL-C serum levels has been estimated as high as 70% in various populations, including Mexican-Americans.^5-8^ Genome wide association studies (GWAS) have successfully identified more than 150 loci associated with lipid levels mainly in European populations,^9-12^ while relatively few GWAS have been performed in Mexicans.^13-16^ The main HDL-C associated loci identified in Europeans are also associated with HDL-C levels in Mexicans, although novel loci have been reported in the latter group.^14^ Notably, a functional variant apparently private to the Americas (*ABCA1* Arg230Cys) was found to be associated with lower HDL-C levels in Mexicans.^17; 18^ Although low HDL-C levels are a well-established cardiovascular risk factor, Mendelian randomization studies have shown that most genetic variants associated with this trait are not associated with cardiovascular risk, suggesting that this relationship is not necessarily causal.^19-21^ In this regard, it has been postulated that pleiotropic effects of the genetic variants or HDL-C particle functionality rather than HDL-C plasma concentrations may affect cardiovascular risk.^22-25^

Low HDL-C levels are highly prevalent in the Mexican population.^26-28^ This population group has been underrepresented in GWAS, and few lipid-associated variants have been tested for coronary artery disease (CAD) risk in Mexicans.^29^ Therefore, we performed a GWAS for HDL-C levels in a cohort of Mexican individuals, using a multiethnic array that includes rare and common genetic variants for Hispanic populations. We then sought to replicate these associations in two independent cohorts: the Genetics of Atherosclerotic Disease (GEA) and the Morbid Obesity Surgery (MOBES) studies, and to test their possible effect on CAD risk. Lastly, we explored the effect of the *SIDT2* gene, a member of a novel family of cholesterol transporters,^30^ on lipid metabolism using existing liver transcriptome data from the Hybrid Mouse Diversity Panel, and the effect of the *SIDT2* Val636Ile variant on the cholesterol analog dehydroergosterol (DHE) uptake *in vitro* using a site-directed mutagenesis approach.

## SUBJECTS AND METHODS

### Study populations

#### Discovery phase cohorts

##### Obesity Research Study for Mexican Children (ORSMEC) and Mexican Adult cohorts

The ORSMEC cohort includes 1,080 school-aged children (6-12 years), 553 with normal-weight and 527 with obesity. The Mexican adult cohort includes 1,073 adults aged 18-82 years, 486 with normal-weight and 587 with obesity. Recruitment strategies, inclusion criteria, anthropometric and biochemical characteristics of both ORSMEC and the Mexican Adult cohorts have been previously described.^31; 32^ Demographic and biochemical characteristics are described in Tables S1 and S2. Briefly, obesity, height and weight were measured following standard protocols and calibrated instruments as previously described.^31^ BMI was calculated as body weight in kilograms divided by the square of height in meters (kg/m^2^). In adults, obesity was defined as a BMI ≥30 kg/m^2^ and normal weight as BMI <25 kg/m^2^ and ≥18.5 kg/m^2^ according to World Health Organization (WHO) criteria.^33^ In children, BMI percentile was calculated using age and sex specific BMI reference data, as recommended by the Centers for Disease Control and Prevention, and obesity was defined as BMI percentile ≥95 and normal weight as BMI percentile >25 and ≤75.^34^

#### Replication phase cohorts

##### Genetics of Atherosclerotic Disease (GEA) cohort

The GEA study was designed to examine the genetic bases of premature CAD in the Mexican population. We included 1,095 adults with premature coronary artery disease and 1,559 individuals recruited as controls without CAD or family history of premature CAD, 413 of which had subclinical atherosclerosis defined by the presence of coronary artery calcification on helical computed axial tomography. Recruitment strategy, inclusion criteria, anthropometric and biochemical characteristics have been previously described.^29^ Demographic and biochemical characteristics are described in Table S3.

##### The Mexican Obesity Surgery (MOBES) cohort

This cohort was designed to study the genetic and metabolomic bases of obesity and metabolic traits in Mexicans and includes 555 individuals with obesity (BMI ≥35kg/m^2^) aged 18 to 59 years who underwent bariatric surgery at the Rubén Leñero General Hospital in Mexico City. Inclusion criteria of this cohort were previously described.^35^ Demographic and biochemical characteristics of this cohort are described in Table S4.

#### Native Americans

This cohort included 302 unrelated Native American individuals (Totonacs and Nahuas from East-central Mexico) aged over 18 years. Inclusion criteria, anthropometric and biochemical characteristics of these individuals have been previously described.^18^ Demographic and biochemical characteristics of these individuals are described in Table S5.

#### Ethics Statement

This study was conducted according to the principles expressed in the Declaration of Helsinki and was approved by the Ethics Committees of participant institutions. All adult participants provided written informed consent prior to inclusion in the study. For children, parents or guardians of each child signed the informed consent and children assented to participate. For Totonac and Nahua participants a translator was used as needed.

#### Biochemical measurements

For all cohorts, blood samples were drawn after 8-12 hours of overnight fasting to determine the serum levels of total cholesterol (TC), triglycerides (TG) and high-density lipoprotein cholesterol (HDL-C) by enzymatic assays as previously described.^31^ Low density lipoprotein-cholesterol (LDL-C) levels were calculated with the equation of Friedewald *et al*.^36^ In the GEA cohort, LDL-C levels were estimated with the equation of Friedewald modified by DeLong *et al*.,^37^ and serum levels of Apolipoprotein A1 and B (ApoA1 and ApoB) were measured by immunonephelometry in a BN Pro Spec nephelometer (Dade Behring Marburg GmBH). In children, low HDL-C levels were defined as HDL-C <40 mg/dL according to the US National Cholesterol Education Program (NCEP) Expert Panel on Cholesterol Levels in Children and Adolescents. For adults low HDL-C levels were defined as HDL-C ≤40 mg/dL for men and ≤50 mg/dL for women according to the US NCEP Adult Treatment Panel III (ATP III).^38^

##### Lipidomic study in the MOBES cohort

Lipidomic analysis was performed using 375 serum samples from MOBES cohort participants with previously described methods.^39; 40^ Briefly, liquid chromatography–electrospray ionization–tandem mass spectrometry (LC–ESI–MS/MS) was used for lipidomic analysis on an Agilent 1290 liquid chromatography system. Liquid chromatography was performed on a Zorbax Eclipse Plus 1.8 µm C18, 50 × 2.1 mm column (Agilent Technologies). The mass spectrometer was operated in dynamic/scheduled multiple reaction monitoring (dMRM) mode. There were 630 unique lipid species belonging to 31 lipid classes identified together with 15 stable isotope or non-physiological lipid standards.

### GWAS and quality control

Genomic DNA was isolated from peripheral white blood cells using standard methods. A total of 2,153 children and adults included in the discovery phase were genotyped using the Multi-Ethnic Genotyping Array (MEGA, Illumina, San Diego, CA, USA), which included >1600k SNPs. This array includes both common and rare variants in Latin American, African, European and Asian populations. Standard quality control (QC) measures were as previously described.^32^ Identity-by-descent (IBD) was estimated using Plink v1.07.^41^ A total of 624,242 SNPs remained after QC measures. After imputation with Beagle,^42^ 865,896 SNPs were included in the final analysis. The quantile-quantile (QQ) plot for the HDL-C GWAS was well calibrated for the null hypothesis (λGC = 1.017), indicating adequate control for confounders (Figure S1).

### Selection of SNPs for the replication analyses

Ten SNPs at 7 loci found to be significantly associated with HDL-C levels in the discovery phase (*P*<1×10^−6^) were selected for replication in two independent cohorts (GEA controls and MOBES). These were the lead SNP of 4 loci (rs9457930 in *LPAL2*, rs983309 in *PPP1R3B*, rs1514661 in *ADAMTS20* and rs1077834 in *LIPC*), and 6 independent SNPs at 3 loci (rs12448528 and rs11508026 in *CETP*, rs9282541 and rs4149310 in *ABCA1*, and rs17120425 and rs10488698 near the *APOA5* cluster). SNPs were defined as independent when LD with other variants within the locus was low (r^2^ and D’ <0.2). Selected SNPs were genotyped using KASP assays (LGC, U.S. http://www.lgcgroup.com). The *SIDT2/* rs17120425 variant was also genotyped in 302 Native Mexicans using a TaqMan assay (ABI Prism 7900HT Sequence Detection System, Applied Biosystems). Call rates exceeded 95% and no discordant genotypes were found in 10% of duplicate samples. No SNPs deviated from Hardy–Weinberg equilibrium in any group (*P*>0.05).

### Mendelian Randomization

In order to test the causal effect of HDL-C levels on CAD we performed Mendelian Randomization (MR) analyses by using HDL-C associated SNPs as an instrument. In MR analyses, genetic variants act as proxies for HDL-C levels in a manner independent of confounders.^43^ We used the inverse-variance weighted (IVW) method^44^ which assumes that all genetic variants satisfy the instrumental variable assumptions (including zero pleiotropy). We also performed MR-Egger regression,^45^ which allow each variant to exhibit pleiotropy. Only SNPs found to be associated with HDL-C in the GEA cohort (n=7) were included in the MR analyses. Both methods were performed with the aid of the Mendelian Randomization R package.^46^

### Ancestry Analysis

Global ancestry was estimated as previously described.^32^ Briefly, European (CEU) and Yoruba (YRI) individuals from the 1000 genomes project and fifteen Nahua and Totonac trios (Native American or NAT) were used as reference populations for ancestry analyses. Multidimensional scaling components were calculated in Plink v1.07.^41^ Ancestral proportions were determined with Admixture.^47^ Local ancestry was determined to identify the origin of chromosomal segments within the 11q23 region using RFMix.^48^

### *Positive selection analysis of* the *SIDT2 Val636Ile variant*

Extended haplotype homozygosity (EHH) values were estimated to seek whether the *SIDT2* derived “A” allele shows evidence of positive selection in a sample of Native Mexican individuals. Sabeti *et al*.^49^ introduced EHH to exploit the decay of haplotype homozygosity as a function of genetic distance from a focal SNP. For this purpose, we used SNP 6.0 microarray data (Affymetrix) from 233 Native Mexicans (Nahuas and Totonacs), and genotyped *SIDT2*/rs17120425 in these individuals. The merged data were phased using Beagle.^42^ Phased alleles were coded as 0/1 (ancestral “G”/derived “A”) to obtain EHH values using the Selscan program.^50^

### HEK293T cell cultures and wildtype and SIDT2/Ile636 transfection

Human embryonic kidney (HEK293T) cells were purchased from the American Type Culture Collection (ATCC, Manassas, VA). Cells were grown on 35 mm culture dishes using Dulbecco’s modified Eagles medium (DMEM) (Invitrogen, Carlsbad, CA) supplemented with 10% of heat inactivated fetal bovine serum (Wisent, premium quality, Canada), penicillin-streptomycin and glutamine (Life Technologies) in an incubator with humidity control at 37 °C and 5% CO_2_.

Human *SIDT2* was cloned from human cDNA CGI-40 (AF151999.1) obtained from the Riken Consortium (Japan). The product was cloned in pEGFP-N1 (Clontech, Mountain View, CA. USA). The SIDT2/Ile636 variant was produced using the QuikChange® Site-Directed Mutagenesis Kit (Stratagene, Santa Clara, CA. USA) following manufacturer’s instructions. The wildtype SIDT2/Val636 variant will be referred to as SIDT2, and the isoleucine variant will be referred as Ile636/SIDT2. The primers used to change valine for isoleucine were the following: FORWARD 5’-GCGTTCTGGATC<u>ATT</u>TTCTCCATCATT-3’ and REVERSE 5’- CGCAAGACCTAG<u>TAA</u>AAGAGGTAGTAA-3’. Constructs were sequenced prior to use.

The plasmids containing SIDT2-GFP and Ile636/SIDT2-GFP were transfected to HEK293T cells grown on 35 mm Petri dishes with a glass bottom (MatTek, Ashland, MA, USA). For transfection we used a mixture of 1 µg of DNA and 6 µl CaCl_2_ reaching 60 µl of final volume with distilled H_2_O. The mixture was added by dropping to HeBS buffer (50mM HEPES, 280mM NaCl, 1.5mM Na_2_HPO_4_, pH 7.05) and the final mixture was incubated for 30 minutes prior to replacing with DMEM. The cells were incubated overnight with the transfection mixture, which was then replaced with fresh medium to perform the assays 24 hours later.

### Fluorescent cholesterol analog dehydroergosterol (DHE) uptake experiments

The naturally occurring blue fluorescent cholesterol analog DHE was purchased from Sigma (Saint Louis, MO). HEK293T cells expressing either SIDT2-GFP or SIDT2/Ile636-GFP were incubated with 5mM DHE solution, adding 300 mM methyl-β-cyclodextrin (MβCD). The solution was carefully resuspended and diluted in PBS to obtain a DHE/MβCD ratio of 1:8 (mol/mol) and 100 µL of this solution were added to the HEK293T cells. Cells were monitored using the scanning confocal microscope (FV1000, Olympus Japan). Focal plane was positioned at the middle of the cells. Confocal images were acquired every 15 seconds with no averaging to reduce photobleaching. Excitation was at 300 nm using a solid-state laser and emission was collected at 535 nm.

### Liver transcriptome analysis in the Hybrid Mouse Diversity Panel (HMDP) and humans

#### HMDP

The HMDP is a collection of approximately 100 well-characterized inbred strains of mice that can be used to analyze the genetic and environmental factors underlying complex traits such as dyslipidemia, obesity, diabetes, atherosclerosis and fatty liver disease. We analyzed the liver transcriptome of strains from this panel carrying the human cholesteryl ester transfer protein (CETP) and the human ApoE3-Leiden transgenes. At the age of about 8 weeks, these mice were placed on a “Western Style” synthetic high fat diet supplemented with 1% cholesterol.^51^ After 16 weeks on this diet, plasma lipid profiles were measured by colorimetric analysis as previously described^52; 53^ and animals were euthanized for the collection of liver tissue. Total RNA was isolated from the left lobe using the Qiagen (Valencia, CA) RNeasy kit (cat# 74104), as described.^54^ Genome wide expression profiles were determined by hybridization to Affymetrix HT-MG_430 PM microarrays. Microarray data were filtered as previously described.^55^ The ComBat method from the SVA Bioconductor package was used to remove known batch effects.^56^ All animal work was conducted according to relevant national and international guidelines and was approved by the UCLA Institutional Animal Care and Use Committee (IACUC).

#### Humans

Total RNA was extracted from liver biopsies of 144 MOBES participants using Trizol reagent (Invitrogen). Clinical and biochemical characteristics of this subgroup of patients are described in Table S6. RNA sequencing was performed as previously described.^57^ Briefly, RNA quality was assessed using the Bioanalyzer RNA chip analysis to ensure that the RNA integrity number was >7. Complementary DNA libraries were prepared using the TruSeq RNA Stranded Total RNA Library Preparation kit (Illumina) and sequenced using an Illumina HiSeq2500 instrument, generating approximately 50 million reads/sample. After data quality control, sequencing reads were mapped to the human reference genome using TopHat software v2.0.1^58^ and quantified using Cufflinks software.^59^

### Statistical Methods

For the discovery phase, genome-wide association with HDL-C was tested independently in 4 groups (normal-weight and obese children and normal-weight and obese adults) under an additive linear mixed model with sex, age and BMI percentile (children) or BMI (adults) as fixed effects, and the genetic relatedness matrix as a random effect. An inverse variance method was used to perform a meta-analysis of the 4 groups.^60^ Genetic relationship matrices from genome-wide data were considered for the analysis using GCTA software.^61^ A *P*-value <1×10^−8^ was considered genome-wide significant, suggestive significance was defined as a *P*-value <1×10^−6^. Group heterogeneity in the meta-analysis was evaluated by I^2^ and Cochrane’s Q^62^ using the R package meta. We used publicly available databases such as the GWAS Catalog (https://www.ebi.ac.uk/gwas/home) to annotate associated SNPs. SNPs within a 1Mb range of the *SIDT2*/Val636Ile variant were included in a locus zoom plot.^63^

For the validation phase, linear regression under additive models was used to test for genetic associations with lipid traits (HDL-C, LDL-C, TC and TG levels) in the GEA control, MOBES and Native American cohorts, and to test for associations with lipid classes in the MOBES cohort. Genetic associations with premature CAD in the GEA cohort were tested using multiple logistic regression under additive models. All tests were adjusted by age, sex and BMI. Associations were tested using the SPSS Statistics package (IBM SPSS Statistics, version 24, Chicago, IL, USA), and statistical significance was considered at *P*<0.05.

Correlations of *SIDT2* liver expression with serum lipid levels and the liver transcriptome were performed using biweight midcorrelation (bicor) coefficient with the R package WGCNA, a robust alternative to Pearson’s correlation coefficient not sensitive to outliers.^64^ Genes significantly correlated with *SIDT2* expression in liver (*P*≤1.0×10^−4^) were tested for pathway enrichment analysis using Metascape and ToppGene Suite software.^65; 66^ Enrichment *P* values <0.05 after FDR correction were considered significant.^67^

## RESULTS

Low HDL-C levels were highly prevalent in Mexican children and adults (38 and 27%, respectively). This trait was significantly more frequent in obese as compared to lean individuals (43.6% vs 12.7% respectively in children, and 81.6% vs 35.4% respectively in adults; *P*<0.001) (Tables S1 and S2). To identify loci associated with HDL-C levels, we carried out a GWAS in 2 independent cohorts of Mexican children and adults using a multi-ethnic array. Of note, the same HDL-C associated loci were found in children and adults, regardless of obesity status, and effect sizes were similar and showed the same directionality. There was no significant evidence of heterogeneity (Table S7), and therefore a fixed-effects meta-analysis was conducted.

In total, we identified 64 variants distributed across 7 *loci* associated with HDL-C levels with genome wide or suggestive significance (*P*<1.0×10^−6^) after adjusting for age, sex, BMI and ancestry (Figure 1, Table S7). Most of the 64 variants were also associated with other lipid parameters (Table S7). Four SNPs showed genome-wide significance (*P*<1.0×10^−8^), rs11508026 and rs12448528 within the *CETP* locus on chromosome 16 (B=3.02 mg/dL; *P=*4.46×10^−18^ and B=-2.79 mg/dL; *P=*5.92×10^−15^, respectively); rs9282541 within the *ABCA1* gene on chromosome 9 (Arg230Cys, B=-3.44 mg/dL; *P*=3.99×10^−13^); and rs17120425 within the *SIDT2* gene (Val636Ile, B=3.31 mg/dL; *P*=1.52×10^−11^), the latter associated with HDL-C levels for the first time (Table 1). The gender stratified meta-analysis showed that the effect size of *SIDT2*/Val636Ile on HDL-C levels was similar in females (B=3.48, *P=*1.9×10^−7^) and males (B=3.27, *P=*1.4×10^−5^) (Table S8). Figure 2 shows the locus zoom plot for the rs17120425 (*SIDT2)* region, including 658 SNPs spanning 1 Mb in chromosome 11q23. No nearby SNPs in LD (r^2^>0.2) with rs17120425 (*SIDT2*) were found.

**Table 1.**
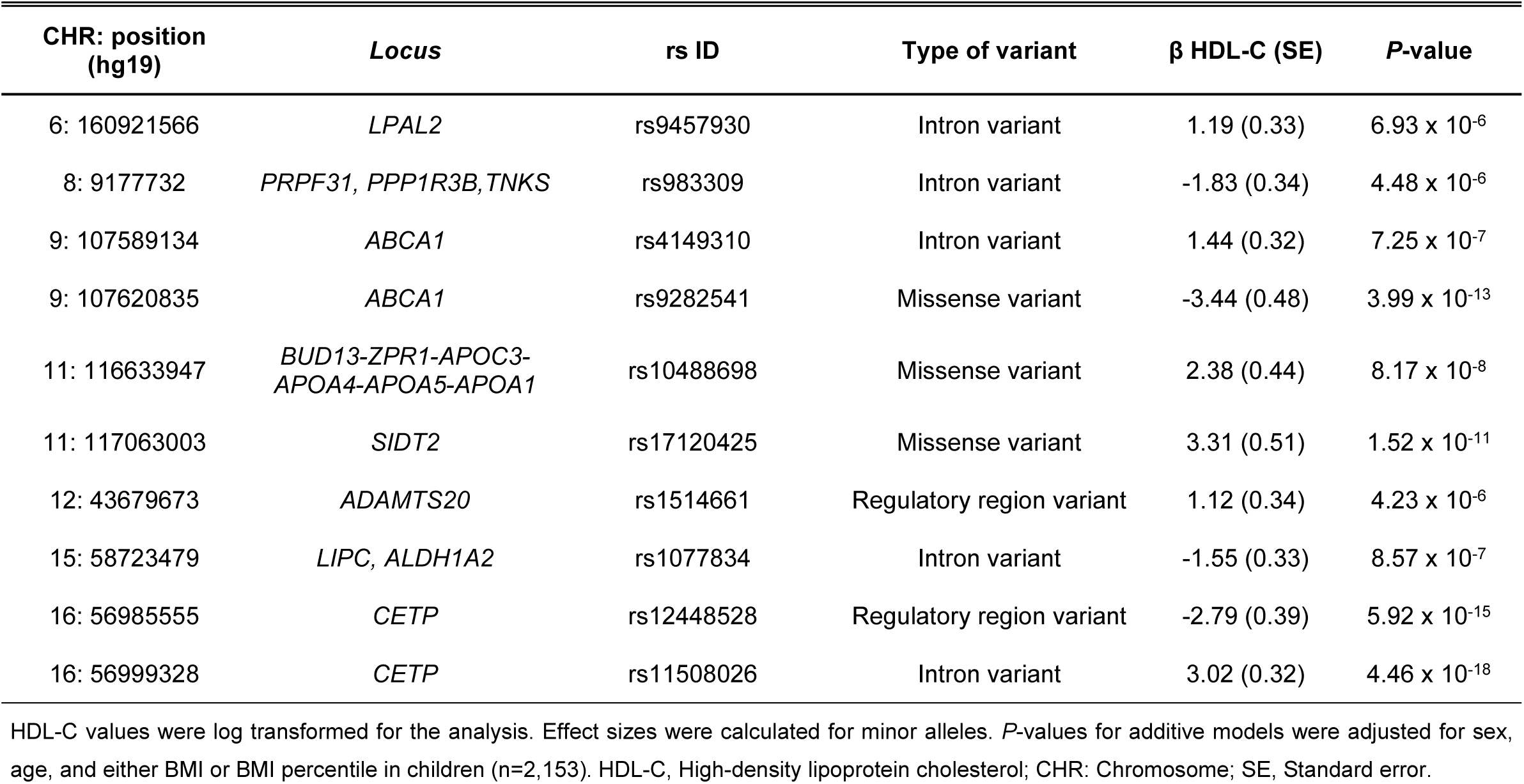
Lead SNPs associated with HDL-C levels in the meta-analysis including Mexican children and adults.

**Figure 1.**
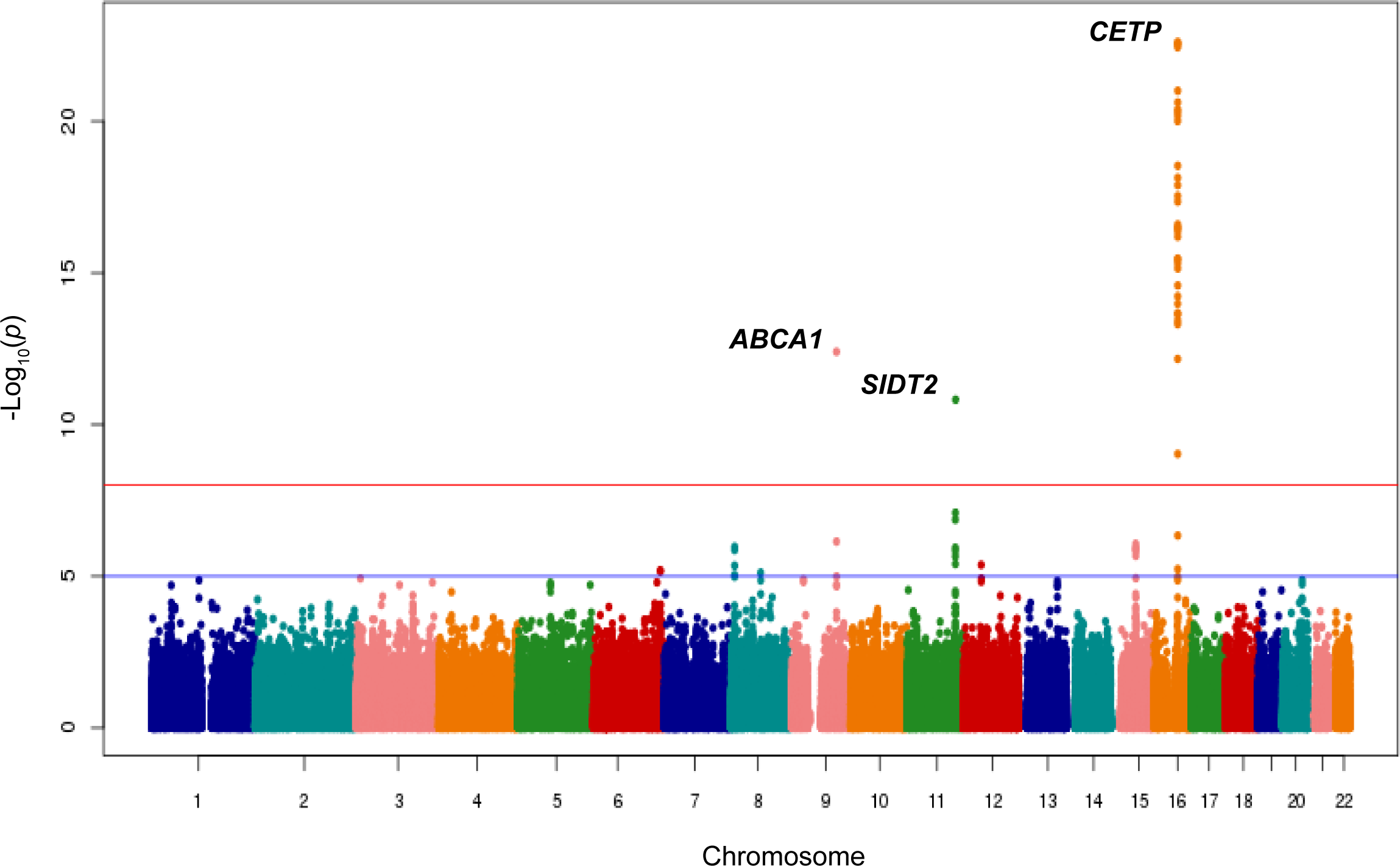
Manhattan plot for HDL-C levels in the discovery phase. Plot showing the -log10 transformed *P*-value of SNPs for 2153 Mexican children and adults. The red line indicates the genome-wide significance level (*P*=5×10^−8^). Genes closest to the SNP with the lowest *P*-value at each locus are indicated.

**Figure 2.**
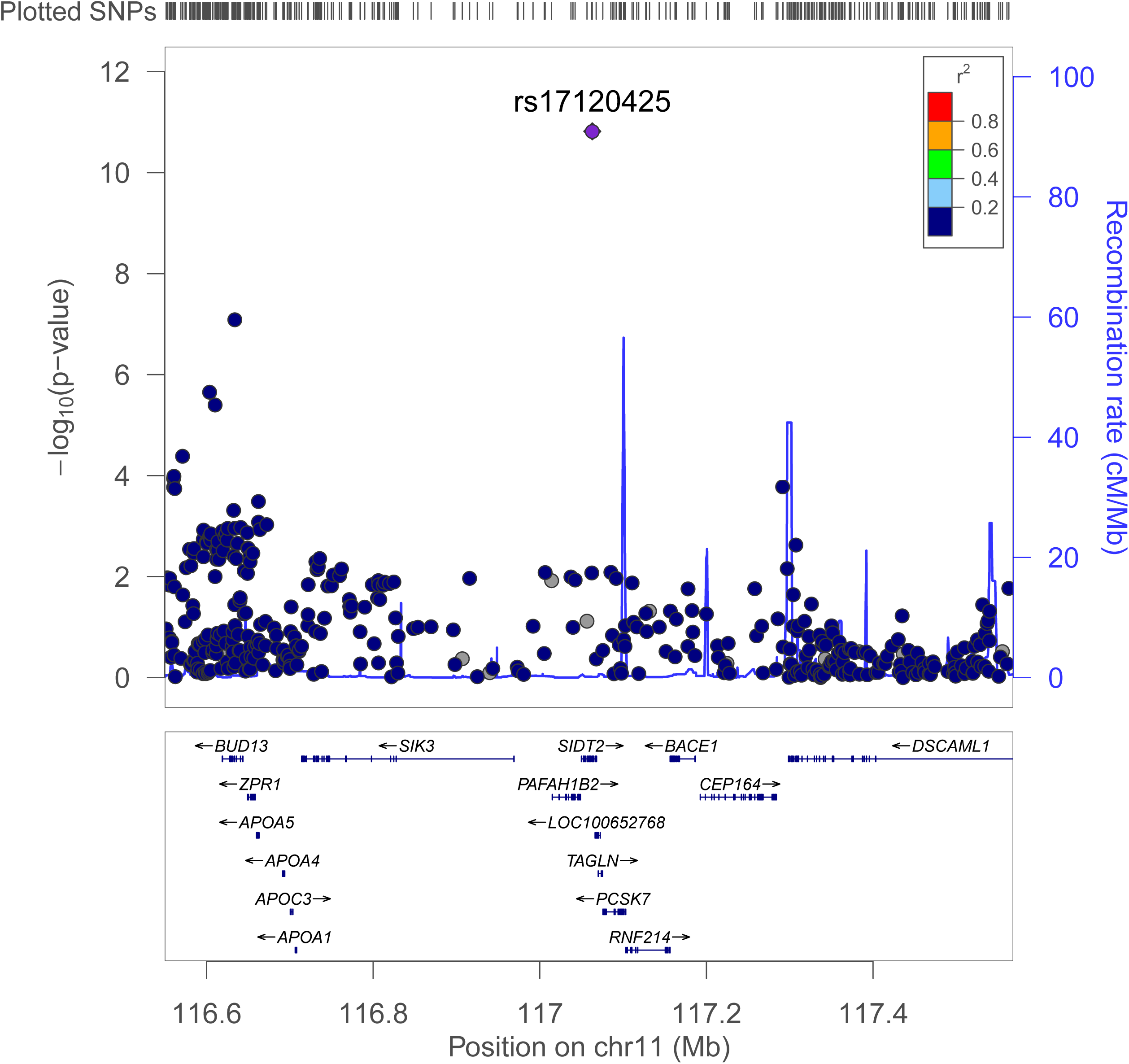
Locus zoom view of variants within the 11q23 region (*SIDT2* locus) associated with HDL-C levels. SNPs are colored based on their correlation (r^2^) with the *SIDT2/*Val636Ile variant (purple diamond), which showed the strongest association with HDL-C levels (*P*=1.5×10^−11^). Arrows on the horizontal blue lines show the direction of transcription, and rectangles represent exons. *P-*values indicate significance of associations found in the discovery phase.

Notably, minor allele frequencies of *ABCA1*/Arg230Cys and *SIDT2*/Val636Ile variants are highest in populations from the Americas (Table S9). We then genotyped the *SIDT2/*Val636Ile variant in two independent Native American populations from central Mexico (Totonacs and Nahuas), which was also associated with higher HDL-C levels in these indigenous groups (B=2.81 mg/dL; *P*=0.027) (Table S10). Moreover, the rs17120425 “A” allele was significantly more frequent in Native Mexicans (15%) than in Mexican Mestizos (10.3%, *P*=0.001) (Table S9), and local ancestry analysis revealed that this allele was found in a block of Native American origin in 98% of individuals. However, according to EHH analysis, LD extension break down was similar in the ancestral and derived rs17120425 alleles, with no evidence of positive selection (Figure S2).

### Replication of associations with HDL-C Levels and other lipid traits in independent cohorts

We then sought to replicate associations with HDL-C levels in 1,559 controls without CAD from the GEA cohort. Seven of the 10 variants associated with HDL-C levels in the discovery phase replicated in GEA controls (*P<*0.010, Table 2): rs12448528 and rs11508026 within the *CETP* gene (*P=*7.2×10^−11^ and *P*=2.9×10^−9^, respectively), rs983309 (*PPP1R3B, P=*4.0×10^−6^), rs17120425/Val636Ile (*SIDT2, P=*7.0×10^−6^), rs9282541 and rs4149310 (*ABCA1, P*=2.0×10^−5^ and *P=*0.01, respectively) and rs10488698 (*BUD13-ZPR1-APOC3-APOA4-APOA5-APOA1* cluster, *P*=6.5×10^−5^). Altogether, these seven variants explained ∼25% of HDL-C level variation (*P*_Genetic Risk Score_=7.0×10^−15^). Table 2 shows associations of these SNPs with other lipid traits in the GEA cohort. *SIDT2/*/Val636Ile and rs10488698 (*BUD13-ZPR1-APOC3-APOA4-APOA5-APOA1* cluster) were both associated with higher HDL-C and ApoA1 levels and lower LDL-C and ApoB levels. SNP rs10488698, but not *SIDT2*/Val636Ile was also significantly associated with lower TG levels. These two SNPs were the only variants associated both with higher ApoA1 and lower ApoB levels. In the MOBES cohort which includes 555 individuals with obesity, rs11508026 (*CETP, P=*9.6×10^−5^), rs9282541 (*ABCA1, P*=1.1×10^−4^) and *SIDT2/*Val636Ile (*P=*0.003) were significantly associated with HDL-C levels (Table 3).

**Table 2.** Association of selected SNPs with lipid traits in the GEA cohort.

| CHR: position<br>(hg19) | Locus | rs ID | HDL-C |  | TC |  | LDL-C |  | TG |  | ApoA1 |  | ApoB |  |
| --- | --- | --- | --- | --- | --- | --- | --- | --- | --- | --- | --- | --- | --- | --- |
|  |  |  | B (SE) | P-value | B (SE) | P-value | B (SE) | P-value | B (SE) | P-value | B (SE) | P-value | B (SE) | P-value |
| 6: 160921566 | *LPAL2* | rs9457930 | 0.59<br>(0.49) | 0.230 | 2.40<br>(1.55) | 0.123 | 1.49<br>(1.31) | 0.322 | 5.32<br>(6.24) | 0.837 | -0.02<br>(1.39) | 0.920 | 1.16<br>(1.15) | 0.286 |
| 8: 9177732 | *PRPF31,*<br>*PPP1R3B, TNKS* | rs983309 | -2.32<br>(0.50) | 4.0x10<sup>-6</sup> | -4.30<br>(1.60) | 0.007 | -2.69<br>(1.35) | 0.016 | 4.03<br>(6.46) | 0.076 | -4.80<br>(1.43) | 0.001 | 0.19<br>(1.19) | 0.982 |
| 9: 107589134 | *ABCA1* | rs4149310 | 1.25<br>(0.49) | 0.010 | 3.68<br>(1.55) | 0.018 | 0.83<br>(1.32) | 0.650 | 14.86<br>(6.22) | 0.011 | 4.22<br>(1.38) | 0.001 | 0.94<br>(1.15) | 0.371 |
| 9: 107620835 | *ABCA1* | rs9282541 | -4.22<br>(0.98) | 2.0x10<sup>-5</sup> | -6.29<br>(2.85) | 0.027 | -2.23<br>(2.53) | 0.622 | 0.68<br>(8.33) | 0.640 | -11.02<br>(3.10) | 5.3x10<sup>-5</sup> | -1.46<br>(2.28) | 0.607 |
| 11: 116633947 | *BUD13-ZPR1-*<br>*APOC3-APOA4-*<br>*APOA5-APOA1* | rs10488698 | 2.75<br>(0.69) | 6.5x10<sup>-5</sup> | -4.17<br>(2.20) | 0.058 | -4.58<br>(1.85) | 0.027 | -16.94<br>(8.83) | 0.048 | 4.65<br>(1.96) | 0.018 | -4.10<br>(1.63) | 0.017 |
| 11: 117063003 | *SIDT2* | rs17120425 | 3.35<br>(0.75) | 7.0x10<sup>-6</sup> | -3.66<br>(2.31) | 0.114 | -6.22<br>(1.96) | 0.004 | -4.75<br>(8.75) | 0.477 | 10.29<br>(2.14) | 2.6x10<sup>-6</sup> | -3.92<br>(1.74) | 0.038 |
| 12: 43679673 | *ADAMTS20* | rs1514661 | -0.12<br>(0.50) | 0.805 | -0.25<br>(1.57) | 0.874 | -0.03<br>(1.32) | 0.773 | 0.43<br>(6.35) | 0.492 | -0.80<br>(1.41) | 0.677 | 0.84<br>(1.16) | 0.381 |
| 15: 58723479 | *LIPC, ALDH1A2* | rs1077834 | -0.59<br>(0.46) | 0.200 | -0.15<br>(1.46) | 0.919 | 1.66<br>(1.23) | 0.090 | -5.44<br>(5.87) | 0.025 | -3.77<br>(1.30) | 0.009 | -0.90<br>(1.08) | 0.281 |
| 16: 56985555 | *CETP* | rs12448528 | -3.77<br>(0.57) | 7.2x10<sup>-11</sup> | -6.02<br>(1.85) | 0.001 | -2.96<br>(1.57) | 0.062 | 3.91<br>(7.48) | 0.340 | -5.26<br>(1.66) | 2.1x10<sup>-4</sup> | -2.09<br>(1.39) | 0.118 |
| 16: 56999328 | *CETP* | rs11508026 | 2.79<br>(0.47) | 2.9x10<sup>-9</sup> | 2.25<br>(1.51) | 0.136 | -0.62<br>(1.28) | 0.830 | 1.38<br>(6.06) | 0.621 | 4.08<br>(1.35) | 0.001 | -0.49<br>(1.12) | 0.979 |

LDL-C, TG, ApoA1 and ApoB levels were log transformed for the analysis. Effect sizes were calculated for the minor alleles. *P*-values for additive models were adjusted for sex, age, and BMI (n=1,559). CHR: Chromosome; MAF, minor allele frequency; HDL-C, High density lipoprotein cholesterol; TC, Total cholesterol; LDL-C, Low density lipoprotein cholesterol; TG, Triglycerides; ApoA1, Apolipoprotein A1; ApoB, Apolipoprotein B.

**Table 3.**
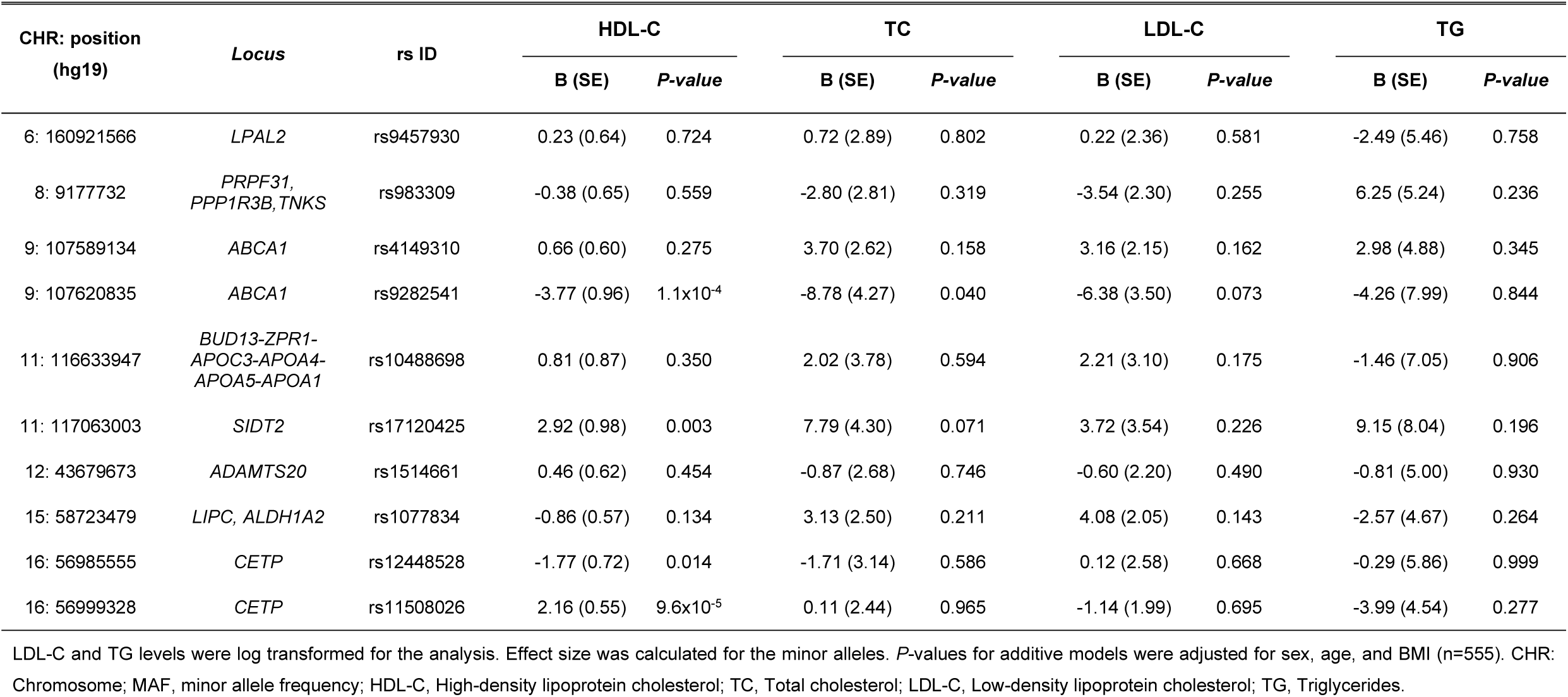
Association of selected SNPs with lipid traits in the MOBES cohort.

| CHR: position<br>(hg19) | Locus | rs ID | HDL-C |  | TC |  | LDL-C |  | TG |  |
| --- | --- | --- | --- | --- | --- | --- | --- | --- | --- | --- |
|  |  |  | B (SE) | P-value | B (SE) | P-value | B (SE) | P-value | B (SE) | P-value |
| 6: 160921566 | *LPAL2* | rs9457930 | 0.23 (0.64) | 0.724 | 0.72 (2.89) | 0.802 | 0.22 (2.36) | 0.581 | -2.49 (5.46) | 0.758 |
| 8: 9177732 | *PRPF31,*<br>*PPP1R3B, TNKS* | rs983309 | -0.38 (0.65) | 0.559 | -2.80 (2.81) | 0.319 | -3.54 (2.30) | 0.255 | 6.25 (5.24) | 0.236 |
| 9: 107589134 | *ABCA1* | rs4149310 | 0.66 (0.60) | 0.275 | 3.70 (2.62) | 0.158 | 3.16 (2.15) | 0.162 | 2.98 (4.88) | 0.345 |
| 9: 107620835 | *ABCA1* | rs9282541 | -3.77 (0.96) | 1.1x10<sup>-4</sup> | -8.78 (4.27) | 0.040 | -6.38 (3.50) | 0.073 | -4.26 (7.99) | 0.844 |
| 11: 116633947 | *BUD13-ZPR1-*<br>*APOC3-APOA4-*<br>*APOA5-APOA1* | rs10488698 | 0.81 (0.87) | 0.350 | 2.02 (3.78) | 0.594 | 2.21 (3.10) | 0.175 | -1.46 (7.05) | 0.906 |
| 11: 117063003 | *SIDT2* | rs17120425 | 2.92 (0.98) | 0.003 | 7.79 (4.30) | 0.071 | 3.72 (3.54) | 0.226 | 9.15 (8.04) | 0.196 |
| 12: 43679673 | *ADAMTS20* | rs1514661 | 0.46 (0.62) | 0.454 | -0.87 (2.68) | 0.746 | -0.60 (2.20) | 0.490 | -0.81 (5.00) | 0.930 |
| 15: 58723479 | *LIPC, ALDH1A2* | rs1077834 | -0.86 (0.57) | 0.134 | 3.13 (2.50) | 0.211 | 4.08 (2.05) | 0.143 | -2.57 (4.67) | 0.264 |
| 16: 56985555 | *CETP* | rs12448528 | -1.77 (0.72) | 0.014 | -1.71 (3.14) | 0.586 | 0.12 (2.58) | 0.668 | -0.29 (5.86) | 0.999 |
| 16: 56999328 | *CETP* | rs11508026 | 2.16 (0.55) | 9.6x10<sup>-5</sup> | 0.11 (2.44) | 0.965 | -1.14 (1.99) | 0.695 | -3.99 (4.54) | 0.277 |

LDL-C and TG levels were log transformed for the analysis. Effect size was calculated for the minor alleles. *P*-values for additive models were adjusted for sex, age, and BMI (n=555). CHR: Chromosome; MAF, minor allele frequency; HDL-C, High-density lipoprotein cholesterol; TC, Total cholesterol; LDL-C, Low-density lipoprotein cholesterol; TG, Triglycerides.

The association of *SIDT2*/Val636Ile with HDL-C levels was highly significant in the conjoint analysis including the discovery phase and replication cohorts (B=3.21, *P*=5.5×10^−21^).

### The SIDT2 Val636Ile variant is associated with serum glycerophospholipid levels in the MOBES cohort

We then sought associations of the 4 HDL-C associated SNPs in the MOBES cohort with lipid classes known to be the main components of HDL-C lipoprotein particles (19 classes of cholesterol esters, phospholipids and TG).^68; 69^ *SIDT2/Val636Ile* was significantly associated with higher glycerophospholipid serum levels (*P*<0.05) (Figure 3), particularly with total phosphatidylcholine (PC), phosphatidylethanolamine (PE), phosphatidylglycerol (PG), phosphatidylinositol (PI) and phosphatidylserine (PS) as adjusted by age, sex, BMI and lipid lowering treatment. In contrast, *ABCA1* (rs9282541) and *CETP* variants (rs12448528 and rs11508026) were not significantly associated with any of these lipid classes.

**Figure 3.**
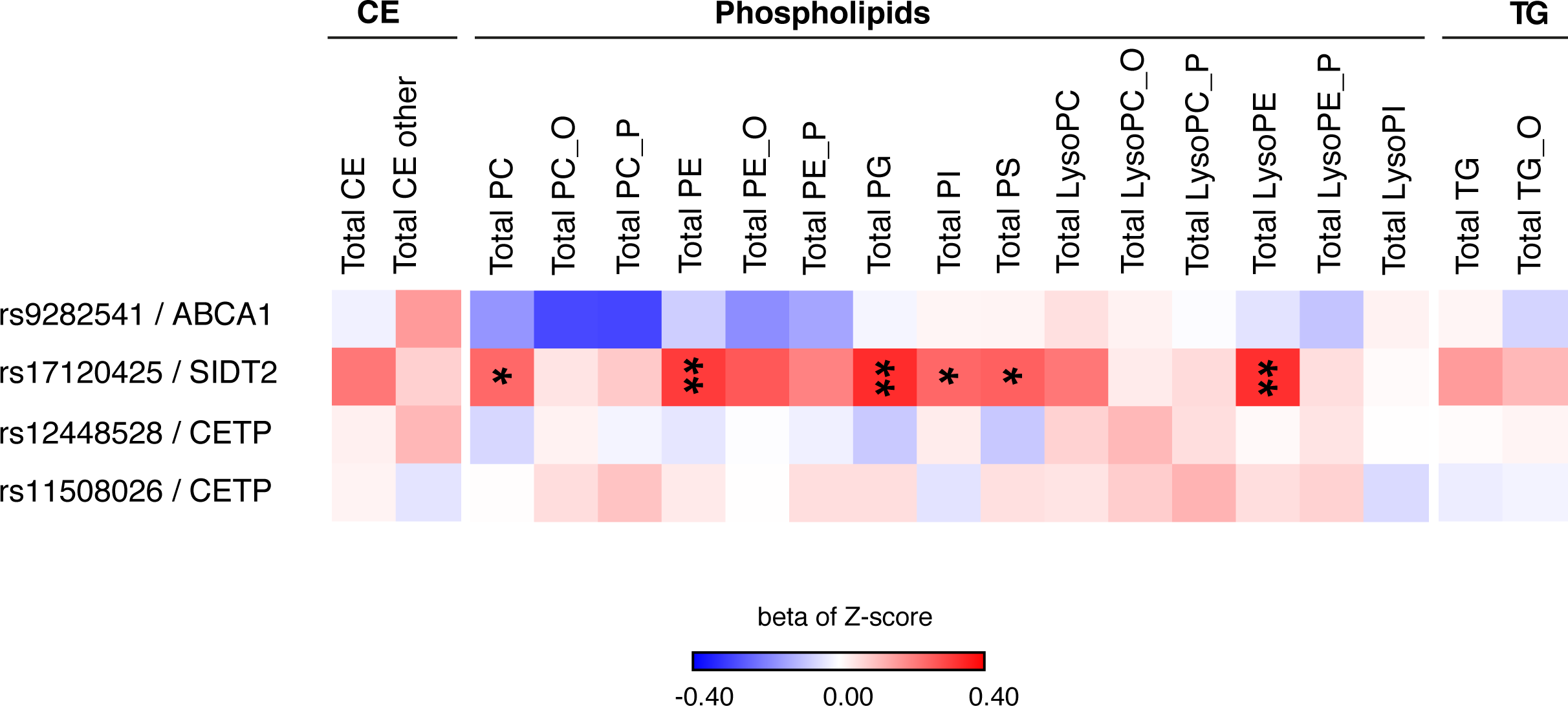
Heat map of associations between the *SIDT2*/Val636Ile variant and lipid classes in the MOBES cohort. Color intensity reflects the Beta value (red for positive, blue for negative) obtained from linear regression between *SIDT2*/Val636Ile and lipid classes in the MOBES study (n=375) adjusted by age, sex and BMI. ^*^*P*-value ≤0.05, ^**^ *P*-value ≤0.001.

### Association with premature CAD and Mendelian randomization

We then tested whether HDL-C variants were also associated with premature CAD in the GEA cohort. Four of the ten variants were significantly associated with CAD risk (rs9282541/*ABCA1*, rs10488698/*APOA1* cluster, rs1077834/*LIPC* and rs17120425/*SIDT2*). Rs10488698/*APOA1-*cluster and *SIDT2/*Val636Ile variants were associated with higher HDL-C levels and lower CAD risk, while rs9282541 (*ABCA1*) and rs1077834 (*LIPC*) were associated with both lower HDL-C levels and lower CAD risk (Table 4). Mendelian randomization analyses were performed including only the 7 variants significantly associated with HDL-C levels in the GEA control cohort. There was no evidence of a causal effect of HDL-C levels on CAD (IVW *P*=0.253 and MR-Egger *P*= 0.509) (Table S11).

**Table 4.** Association of HDL-C associated SNPs with premature coronary artery disease in the GEA cohort.

| CHR: position<br>(hg19) | Locus | rs ID | MAF |  | OR (95% CI) | P-value |
| --- | --- | --- | --- | --- | --- | --- |
|  |  |  | Controls<br>(n=1,559) | Premature CAD<br>(n=1,095) |  |  |
| 6: 160921566 | *LPAL2* | rs9457930 | 37.9 | 37.0 | 0.98 (0.86-1.11) | 0.704 |
| 8: 9177732 | *PRPF31,*<br>*PPP1R3B, TNKS* | rs983309 | 29.3 | 30.5 | 1.06 (0.93-1.21) | 0.370 |
| 9: 107589134 | *ABCA1* | rs4149310 | 34.4 | 36.6 | 1.11 (0.98-1.26) | 0.116 |
| 9: 107620835 | *ABCA1* | rs9282541 | 10.4 | 4.8 | 0.44 (0.30-0.65) | 3.9x10<sup>-5</sup> |
| 11: 116633947 | *BUD13-ZPR1-APOC3-*<br>*APOA4-APOA5-APOA1* | rs10488698 | 12.9 | 10.7 | 0.80 (0.67-0.97) | 0.023 |
| 11: 117063003 | *SIDT2* | rs17120425 | 9.5 | 8.0 | 0.80 (0.65-0.99) | 0.039 |
| 12: 43679673 | *ADAMTS20* | rs1514661 | 31.1 | 30.7 | 0.98 (0.87-1.12) | 0.803 |
| 15: 58723479 | *LIPC, ALDH1A2* | rs1077834 | 42.6 | 39.1 | 0.87 (0.77-0.98) | 0.027 |
| 16: 56985555 | *CETP* | rs12448528 | 20.5 | 18.1 | 0.87 (0.75-1.02) | 0.087 |
| 16: 56999328 | *CETP* | rs11508026 | 36.3 | 39.0 | 1.11 (0.99-1.26) | 0.087 |

Associations with premature CAD were adjusted for age, sex and BMI. *P*-values were calculated by logistic regression. HDL-C, High-density lipoprotein cholesterol; CHR: Chromosome; MAF, minor allele frequency; CAD, coronary artery disease; OR, Odds ratio; CI, confidence interval.

### Fluorescent cholesterol analog uptake is enhanced in cells expressing Ile636/SIDT2

It was previously suggested that the transmembrane conserved cholesterol binding (CRAC) domain of murine Sidt2 associates with the cholesterol analogue dehydroergosterol in HEK293 cells.^30^ Because the *SIDT2*/Val636Ile variant is near the transmembrane CRAC domain in the human SIDT2 protein, we evaluated the effect of this variant on DHE uptake in HEK293T cells. Interestingly, DHE uptake was enhanced in cells expressing the Ile636/SIDT2 protein as compared to cells expressing wildtype SIDT2, reaching the highest difference approximately 1.5 minutes after adding DHE to the culture (6.85 ± 0.42 AU vs 9.40 ± 0.73 AU, *P*<0.01) (Figure 4, Video S1).

**Figure 4.**
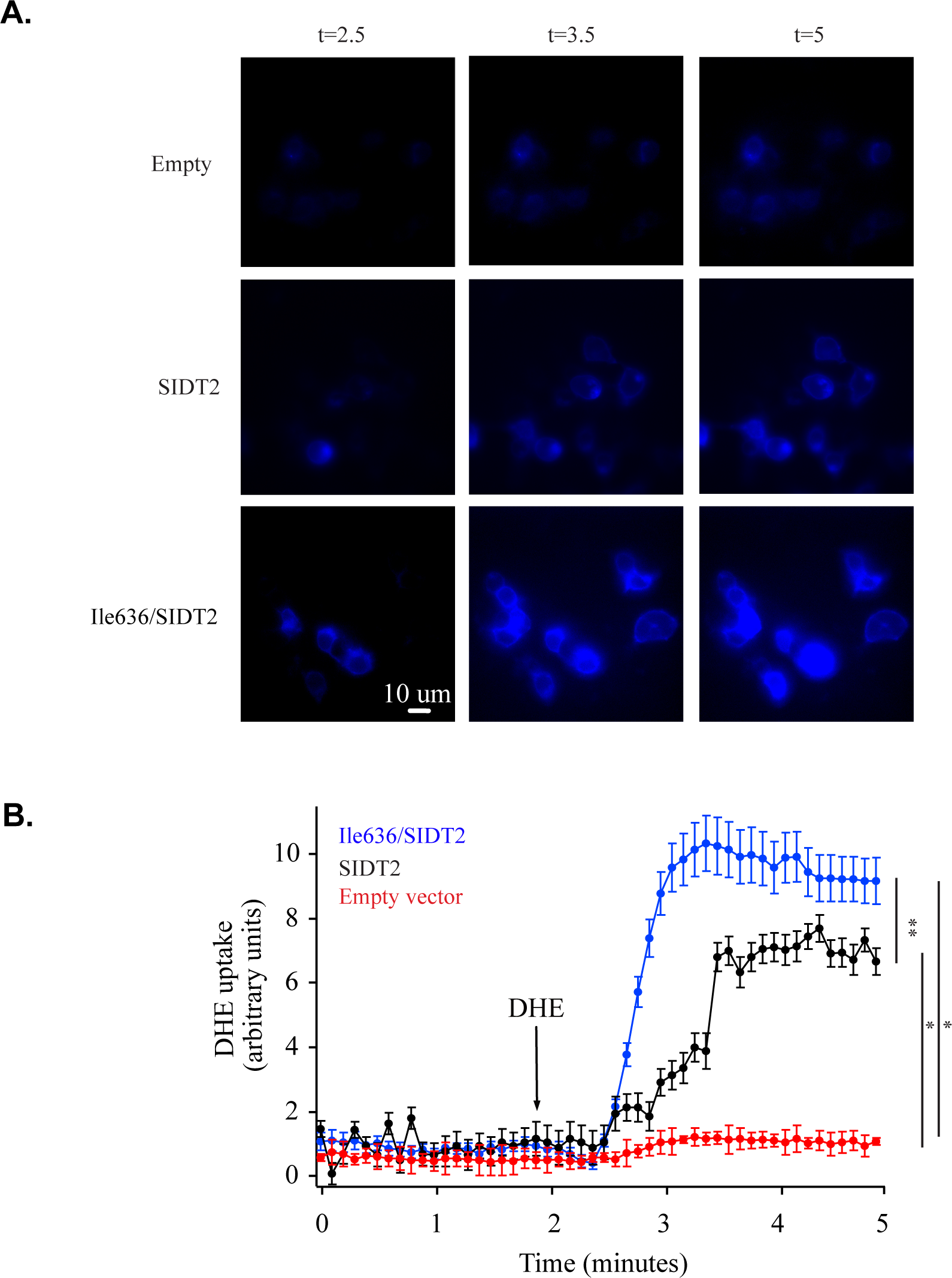
Uptake of the blue fluorescent cholesterol analog dehydroergosterol (DHE) by cells expressing wildtype and Ile636/SIDT2. (A) Confocal microscopy images of HEK293T cells expressing the empty vector, wildtype and Ile636/SIDT2, at times 2.5, 3.5 and 5 minutes after adding the fluorescent cholesterol analog DHE to the culture. (B) Plot showing mean fluorescence intensity over time after adding DHE to the culture. Values show mean ± standard deviations from at least 4 independent experiments, each experiment shows mean values from all the cells in the focal plane. The red dots represent cells transfected with the empty vector; the black dots represent cells expressing wildtype SIDT2, and blue dots represent cells expressing Ile636/SIDT2. Addition of DHE is indicated with an arrow. ^*^*P*<0.05; ^**^*P*<0.01.

### SIDT2 liver expression correlates with the expression of genes involved in lipid and lipoprotein metabolism

In the MOBES cohort, hepatic *SIDT2* expression did not differ according to the presence of the Val636Ile variant (*P=*0.486). Moreover, human *SIDT2* liver expression showed no significant correlation with lipid traits including HDL-C, TC, or TG levels (Figure 5A). In contrast, in HMDP mice fed with an atherogenic diet, hepatic *Sidt2* expression correlated positively with HDL-C levels (r=0.312, *P*=0.002), and negatively with total cholesterol and TG levels (r=-0.381, *P*=1.2×10^−4^; r=-0.304, *P*=0.002, respectively) (Figure 5B).

**Figure 5.**
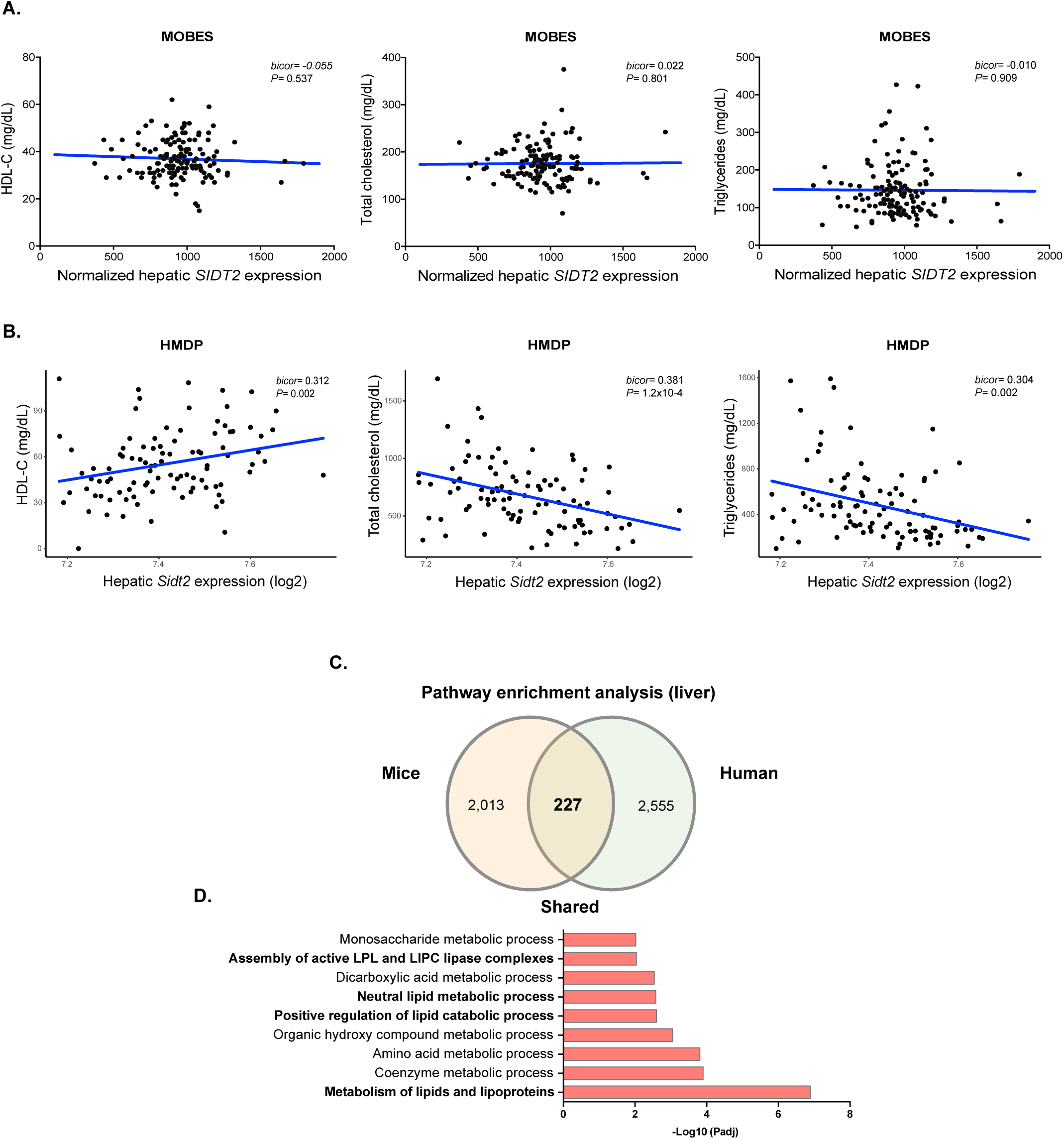
Liver *SIDT2* expression and lipids in mice and humans. Correlations of liver *Sidt2* expression with HDL-C, total cholesterol, and TG serum levels in MOBES cohort participants (A) and mice from the HMDP (B). (C) Venn diagram depicting the overlap of genes significantly correlated with *SIDT2* liver expression in HMDP mice and MOBES cohort participants (*P<*1.0×10^−8^). (D) Significantly enriched pathways in mice and humans.

To analyze correlations between *SIDT2* expression and the liver transcriptome in mice, we used data from the genetically diverse HMDP where environmental factors are controlled. A total of 2,240 genes showed significant correlations with *Sidt2* expression, and the most significantly enriched pathway was lipid and lipoprotein metabolism (*PFDR*=1.5×10^−7^). In human liver tissue *SIDT2* expression correlated with the expression of 2,782 genes. Consistent with observations in mice, the lipid and lipoprotein metabolism pathways were among the most significantly enriched. *SIDT2* expression correlated with the expression of 227 genes that were shared by both mice and humans, and as expected metabolism of lipids and lipoproteins was the most significantly enriched pathway (Figure 5C-D).

## DISCUSSION

Low HDL-C levels are the most common dyslipidemia in Mexicans, both in adults and children.^26; 28^ Consistently, low HDL-C levels were highly prevalent in children and adults of the present study. Moreover, the prevalence of low HDL-C levels was significantly higher in obese than in lean individuals, in line with the high prevalence of metabolic syndrome observed in the Mexican population.^70^

Here, using a GWAS for HDL-C levels in different cohorts of the Mexican population, we identified three loci associated with HDL-C levels at chromosomes 9, 11 and 16. Of note, the effect and direction of the associations were consistent in the 4 study groups (normal weight and obese children and adults). The most significant signal corresponds to the *CETP* locus, which is known to be one of the main drivers of HDL-C levels across populations.^10; 15; 71^ The signal in chromosome 9 is the *ABCA1*/Arg230Cys variant (rs9282541) previously reported as associated with lower HDL-C levels by our group, apparently private to Native American and derived populations.^17; 18^ The third genome-wide significant signal corresponds to a missense variant within the *SIDT2* gene (Val636Ile, rs17120425), associated with higher HDL-C levels. This association was replicated in 3 independent cohorts including Native Americans from Mexico. This is relevant because Native Americans live in rural areas, while the GEA and MOBES cohorts are urban populations, known to have different dietary habits, which may affect HDL-C levels.^72^

To our knowledge, this is the first time that the *SIDT2*/Val636Ile variant has been associated with HDL-C levels. However, intronic *SIDT2* variants have been previously associated with lipid traits and the metabolic syndrome in multi-ethnic cohorts, mainly in Koreans.^73; 74^ Nevertheless, these variants (rs2269399, rs7107152, rs1242229 and rs1784042) are not in LD with *SIDT2*/Val636Ile (r^2^<0.2). Although the *SIDT2*/Val636Ile variant is not private to the Americas, it is more frequent in Native Americans (15%) and Hispanics (6%), than other ethnic groups (internationalgenome.org). Local ancestry analyses revealed that the derived “A” allele was of Native American origin in most individuals. However, the extended haplotype homozygosity (EHH) analysis of *SIDT2*/Val636Ile did not show evidence of recent positive selection.

It is likely that previous GWAS in Mexicans failed to identify this SNP as associated with HDL-C because this variant was not included in the microarray platforms used in these studies and the paucity of Native American references for imputation.^13-16^ This 11q23 region contains several lipid-associated signals, and it is thus necessary to define whether these signals are independent of *SIDT2*/Val636Ile. The *BUD13-ZPR1-APOC3-APOA4-APOA5-APOA1* cluster associated with TG levels is 400 Kb upstream *SIDT2*. Our regional LD analysis demonstrated the lead TG-associated SNP rs964184 (*APOA5*)^9; 13^ and all other SNPs analyzed within this cluster were in low LD with *SIDT2*/Val636Ile (r^2^<0.2). This indicates that the *SIDT2* signal associated with HDL-C and SNPs within the *BUD13-ZPR1-APOC3-APOA4-APOA5-APOA1* cluster are in fact independent. Moreover, a previous GWAS in the Mexican population identified two intronic *SIK3* gene variants within the 11q23 region, rs139961185 associated with TG levels and rs11216230 with higher HDL-C levels.^14^ The latter association was replicated in an independent Hispanic population.^16^ Of note, rs11216230 is more frequent in Mexicans (11%) than in Europeans (1%) and is in high LD with *SIDT2*/Val636Ile in Mexican Americans from the 1000 Genomes project (r^2^=0.75). This suggests that the association observed by Ko *et al*.^14^ could be driven by the *SIDT2*/Val636Ile variant.

In Mendelian randomization analyses, genetic variants act as proxies for HDL-C levels in a manner independent of confounders to analyze the causality of HDL-C levels on coronary artery disease. Our MR analysis is consistent with previous studies suggesting that higher HDL-C levels are not causally protective against coronary heart disease.^19-21^ This suggests that the effect of individual variants on CAD risk may be mediated by pleiotropic effects on other cardiovascular risk factors, or on HDL-C composition and functionality.^22-25; 75^ The *SIDT2*/Val636Ile variant was associated with higher HDL-C levels and with lower risk of premature CAD. We thus explored whether this variant affects other cardiovascular risk parameters in addition to HDL-C levels. Notably, *SIDT2*/Val636Ile was also associated with higher ApoA1 levels, and lower LDL-C and ApoB serum levels in the GEA cohort. APOA1 is the major protein component of HDL-C particles, and a Mendelian randomization analysis in Finnish individuals reported that ApoA1 was not associated with risk of CAD.^76^ A multivariable Mendelian randomization study examining serum lipid and apolipoprotein levels reported that only ApoB retained a robust relationship with the risk of CAD,^77^ and recent Mendelian randomization studies suggest that ApoB is the primary lipid determinant of cardiovascular disease risk.^78; 79^ Thus, the association of *SIDT2*/Val636Ile with decreased cardiovascular risk could be mediated by its effect on ApoB levels.

HDL lipidome composition has been associated with HDL-C functional properties,^80-82^ Notably, of the 4 main variants associated with HDL-C levels in the present study, only *SIDT2*/Val636Ile was associated with lipid species, specifically with higher serum concentrations of several glycerophospholipid classes including PE, PG, PC, PI and PS. It has been reported that HDL-C particles enriched in phospholipids can increase HDL-C stability,^83^ while decreased levels of phospholipids in HDL-C were found to impair cholesterol efflux and decrease the cardiovascular protective effects of HDLs.^69; 82; 84^ Particularly, recent studies indicate that phosphatidylserine, a minor component of the monolayer surface of HDL-C, is enriched in small, dense HDL-C particles, which display potent anti-atherosclerotic activities.^83; 85^ Although we measured phospholipids in serum and not directly in HDL-C particles, a limitation of the study, the association of *SIDT2*/Val636Ile with higher phospholipid levels is consistent with the lower cardiovascular risk conferred by this variant.

The SIDT2 protein is found mainly in lysosome membranes,^86^ is a lysosomal nucleic acid transporter, and is expressed in several tissues, including the liver.^87-91^ The mammalian SIDT2 protein has high homology to the *C. elegans* cholesterol uptake protein-1 (CUP-1).^92^ SIDT2 has been identified as a sterol-interacting protein^93^ and more recently as a cholesterol-binding protein.^30^ Moreover, SIDT2 is predicted to contain two CRAC domains (Cholesterol Recognition/interaction Amino Acid Consensus), found in a broad range of proteins involved in cholesterol transport, metabolism and regulation.^30; 94^ Specifically, the transmembrane CRAC domain from human SIDT1 and mouse SIDT2 appears to bind cholesterol.^30^ The Val636Ile variant and the CRAC domain are located within the same transmembrane segment, and this variant is 19 amino acids upstream the tyrosine CRAC domain residue, suggested to interact with the cholesterol OH-polar group.^30^ It is unknown if the Val636Ile variant modifies the interactions of the CRAC domain with cholesterol, thus affecting circulating lipid levels.

In the present study, HEK293T cells expressing the Ile636/SIDT2 protein showed higher cholesterol analog DHE uptake than those expressing the wildtype protein. This increased uptake observed *in vitro*, may affect circulating levels of cholesterol-rich lipoproteins. Although the mechanism by which this variant increases HDL-C serum levels is unknown, ABCA1-mediated cholesterol efflux and HDL-C formation are primarily dependent on autophagy for cholesterol source.^95^ Because *Sidt2*^*-/-*^ deficient mice show blocked autophagosome maturation and altered hepatic lipid homoeostasis,^96^ it is tempting to speculate that the putative increased function of the Ile636 SIDT2 protein may enhance autophagy-mediated cholesterol flux, and thus ABCA1-mediated HDL-C formation.

*Sidt2* knockout mice show a wide range of metabolic phenotypes including impaired glucose tolerance likely due to compromised NAADP-involved insulin secretion.^96-100^ Meng *et al*.,^100^ showed that *Sidt2* deficient mice present significantly increased serum total cholesterol, TG and LDL-C levels, and significantly lower HDL-C serum levels as compared to *Sidt2*^+/+^ mice. These findings are consistent with the inverse correlation between hepatic *Sidt2* expression and total cholesterol and triglycerides levels, and the direct correlation of *Sidt2* expression with HDL-C levels observed in the HMDP. Moreover, *Sidt2*^-/-^ mice not only have altered lipid serum levels, but also showed impaired liver function and liver steatosis.^96; 99; 100^ Consistently, *Sidt2* expression correlated with increased fat liver content in HMDP mice (data not shown). In contrast, in the MOBES cohort *SIDT2* liver expression did not correlate with serum lipid levels or liver steatosis, and the *SIDT2*/Val636Ile variant showed no association with non-alcoholic fatty liver disease in the GEA or MOBES cohorts (data not shown). Because MOBES cohort participants had morbid obesity and do not represent the overall population, the lack of correlation between *SIDT2* liver expression with lipid levels in this cohort must be interpreted with caution. Despite these differences between mice and humans, *SIDT2* expression correlations with the liver transcriptome revealed that the most significantly enriched pathways were lipid and lipoprotein metabolism in both species. Altogether, these findings support a role of SIDT2 in lipid metabolism. Further studies are required to better understand the mechanisms by which SIDT2 participates in cholesterol and lipoprotein metabolism at the cellular and systemic levels, and its role in cardiovascular risk.

In conclusion, this is the first study assessing genetic variants contributing to HDL-C levels and coronary artery disease in the Mexican population. Our GWAS revealed for the first time that the *SIDT2*/Val636Ile variant is associated with increased HDL-C and phospholipid levels and decreased risk of CAD. We also provide evidence that the *SIDT2*/Val636Ile variant is functional, increasing the uptake of a cholesterol analog *in vitro*. Our data support a role of SIDT2 in cholesterol and lipid metabolism. The mechanisms by which this protein affects lipid and metabolic parameters in humans require further investigation.

## Data Availability

The datasets supporting the current study have not been deposited in a public repository because they are part of other studies in progress, but are available from the corresponding authors on reasonable request.

## Supplemental Data

Supplemental Data include two figures, eleven tables and one video.

## Declaration of Interests

The authors declare no competing interests.

## Acknowledgements

We thank Luz E. Guillén for her technical support. We also thank Productos Medix S.A. de C.V. for their support to perform this study. This work was supported by grants FOSISS-289699 and PEI-230129 from Mexican National Council for Science and Technology (CONACyT).

## Web Resources

R, https://www.r-project.org

PLINK, http://zzz.bwh.harvard.edu/plink

gnomAD, https://gnomad.broadinstitute.org

1000 Genomes, https://www.internationalgenome.org

GWAS Catalog, https://www.ebi.ac.uk/gwas/home

WCGNA,https://horvath.genetics.ucla.edu/html/CoexpressionNetwork/Rpackages/WGCNA

ToppGene, https://toppgene.cchmc.org

Metascape, https://metascape.org

OMIM, http://www.omim.org

HGNC, http://www.genenames.org

## FIGURE TITLES AND LEGENDS

**Video S1. Uptake of the fluorescent cholesterol analog dehydroergosterol (DHE) in HEK293T cells expressing wildtype or Ile636/SIDT2.** Representative experiments illustrating cells in the focal plane. Fluorescence was monitored in time using confocal microscopy. DHE uptake in cells expressing the empty vector (upper panel), wildtype SIDT2 and the Ile636/SIDT2 proteins (lower panels).

## Notes

### Competing Interest Statement

The authors have declared no competing interest.

